# Attenuated alpha oscillations as an index of cortical hyperexcitability: Evidence from migraine patients

**DOI:** 10.1101/2021.04.21.21255858

**Authors:** Chun Yuen Fong, Wai Him Crystal Law, Johannes Jacobus Fahrenfort, Jason J. Braithwaite, Ali Mazaheri

## Abstract

Anomalous phantom visual perceptions coupled to an aversion to some visual patterns has been associated with aberrant cortical hyperexcitability in migraine patients. Previous literature has found fluctuations of alpha oscillation (8-14 Hz) over the visual cortex to be associated with the gating of the visual stream. In the current study, we examined whether alpha activity was differentially modulated in migraineurs in anticipation of an upcoming stimulus as well as post-stimulus periods. We used EEG to examine the brain activity in a group of 28 migraineurs (17 with aura/11 without) and 29 non-migraineurs and compared the modulations of alpha power in the pre/post-stimulus period relative to onset of stripped gratings of 3 spatial frequencies 0.5, 3, and 13 cycles per degree (cpd). Overall, we found that that migraineurs had significantly less alpha power prior to the onset of the stimulus relative to controls. Moreover, relative to the control group, migraineurs had significantly greater post-stimulus alpha suppression (i.e event-related desynchronization) induced by the 3 cpd grating at the 2^nd^ half of the experiment, the stimulus most often reported to induce visual disturbances. These findings taken together provide strong support of the presence of elevated cortical excitability in the visual cortex of migraine sufferers. We speculate that cortical hyperexcitation could be the consequence of impaired perceptual learning driven by the dysfunction of GABAergic inhibitory mechanism.

## 1. Introduction

Previous studies have proposed that migraine patients have an elevated visual cortical excitability (often referred to as hyperexcitability) which could account for their (i) interictal experiences of photophobia, (ii) pattern aversion, and (iii) aberrant visual experiences which include distortions and hallucinations / aura (Bouloche, Denuelle, Payoux et al., 2010; Palmer, Chronicle, Rolan et al., 2000; Fong, Law, Braithwaite et al., 2020; Haigh, Karanovic, Wilkinson et al., 2012; van der Kamp, VanDenBrink, Ferrari et al., 1996). The concept of increased hyperexcitability is often seen as an outcome of cortical under-inhibition (Boulloche et al., 2010; McKendrick, Chan, Vingrys et al., 2018; Mulleners, Chronicle Palmer et al., 2001). This abnormality disrupts the balance between cortical inhibition and excitation which weaken the suppressive function of the sensory system, leading to hypersensitivity to light or some visual patterns (e.g. gratings of a specific spatial frequency) by the over-stimulation of localised neural systems.

In the current electroencephalography study, we used modulation of the ongoing alpha activity (8-12 Hz) induced by the onset of visual stimuli to gauge the excitability of the visual cortex of migraine patients. We focused our investigation on modulations of oscillatory activity in the alpha range given that previous work has found support of this rhythm to be involved in gating of visual input via the GABAergic interneuron system (Klimesch, Sauseng, & Hanslmayr, 2007, Jensen & Mazaheri, 2010, Van Diepen, Foxe, & Mazaheri 2019).

The majority of the previous literature looking for aberrant patterns of oscillatory activity in migraine patients has primarily focused on task-free resting state EEG, with often contradictory findings. For example, one study observed interictal patients to have increased theta and delta resting-state activities (Bjork, Stovner, Engstrom et al., 2009) while in another study, migraine patients appeared to show reduced theta, alpha, beta power (Cao, Lin, Chuang et al., 2016). In a recent study, migraineurs were found to have increased resting alpha before and after a visual discrimination task (O’Hare, Menchinelli, & Durrant, 2018). To the best of our knowledge there have been few, if any, studies systematically looking at induced oscillatory changes in the EEG activity of migraine patients in response to visual stimuli.

Evidence for cortical hyperexcitability of migraine sufferers is supported by the observation that they often demonstrate a significantly lower phosphene induction threshold when their visual cortex is stimulated by transcranial magnetic stimulation (TMS: Aurora, Cao, Bowyer, & Welch, 1999; Aurora, Welch, & Al-Sayed, 2003; van der Kamp, VanDenBrink, Ferrari, & van Dijk, 1996; Fumal, Bohotin, Vandenheede, & Schoenen, 2003). Migraine sufferers have also been reported to be less influenced by metacontrast masking effect (Palmer, Chronicle, Rolan et al., 2000) as well as having a lower contrast threshold to certain visual patterns (Haigh, Karanovic, Wilkinson et al., 2012). The predisposition to experience visual discomfort by viewing stripped grating at spatial frequency around 2 to 4 cycles per degree (cpd) for migraineurs, namely pattern glare, are thought to be associated with their cortical hyperexcitability (Fong, Takahashi, & Braithwaite, 2019; Wilkins, 1995; Wilkins, Nimmo-Smith, Tait, et al. 1984). These behavioural findings are in line with electrophysiological evidence, where migraineurs showed enhanced visual evoked potentials (VEPs) to aversive patterns (Oelkers, Grosser, Lang et al., 1999; Fong, Law, Braithwaite et al., 2020).

Further evidence suggesting anomalies in the excitability of the visual cortex of migraine sufferers is the seeming lack of habituation to repetitive visual stimulation. In the typical non-migraine population, repeated visual stimuli evoked reduced event-related potentials suggesting stimulus habituation (Mazaheri & Picton, 2005). This phenomenon is part of a perceptual learning mechanism and may prevent excessive neuronal stress generated at the sensory cortex (Sappey-Marinier, Calabrese, Fein et al., 1992). However, patients suffering from migraine have shown an enhancement of VEPs after a succession of visual stimulation (Schoenen, Wang, Albert et al., 1995). This “potentiation” effect, in the visual cortex of migraineurs can also be observed in other sensory modalities (Coppola, Pierelli, & Schoenen, 2008).

In the current investigation, in order to gain a better understanding of the responsiveness of the visual cortex of migraine patients, we examined the brain activity in a group of 28 migraineurs (17 with aura/11 without) and 29 non-migraineurs. We set out to find support for the cortical hyperexcitability hypothesis by comparing the modulations of alpha power induced by stripped patterns of low, medium and high spatial frequencies (i.e 0.5, 3, and 13 cpd). We focused on the period in anticipation of a visual stimulus (i.e post-cue to pre-stimulus) as well as post-stimulus modulations of alpha activity for the different stimuli. The pre-stimulus level of alpha activity allowed us to gauge the baseline excitability of the visual cortex expecting the arrival of an upcoming stimulus, whereas the post-stimulus alpha modulation gave us an insight into the resources allocated to the processing of the visual stimuli.

## 2. Methods

### 2.1 Participants

Our experiment included 28 self-reported female migraineurs (mean age = 20.9) and 29 healthy female control (mean age = 19.4) with normal/corrected to normal visual acuity (20/25 or better). The participants were all part of a previously published study (Fong et al., 2020). All healthy participants have reported no history of migraine nor any neurological and psychiatric conditions. Amongst the 28 migraine patients, 17 of them were categorised as migraine with aura and 11 as migraine without aura according to the criteria of International Headache Society (Olesen, 2018). The migraine patients in the current study were not regularly taking any prophylactic medications (and had not taken any within 2 weeks of the experiment), nor had chronic migraine, motor migraine aura symptoms or any other comorbid neurological or psychiatric conditions. The EEG sessions were taken at the interictal period of the migraineurs (no migraine attack before 1 week and after at least 2 weeks of the recordings). This study has been approved by the Ethics Committees of the University of Birmingham (reference number: ERNE_14–0875AP1A).

### 2.2 Stimuli, apparatus and trial sequences

Achromatic gratings with Michelson contrast of 0.70 (cd/m2) in three different spatial frequencies (0.5, 3 and 13 cpd, also named as LF, MF, and HF) were used as visual stimuli in the current study. The stimuli were presented at the centre of a 20-inch Dell P2210 LCD computer screen (60 Hz refresh rate and 1680×1050 pixels screen resolution) using E-prime v2.0 software, with a background luminance of 20 cd/m^2^ in free-viewing condition. The stimuli all had an identical ellipse shape with the maximum height x width of 140 mm x 180 mm which gave a visual angle of 9.93×12.68° when the viewing distance was fixed at 80 cm.

Every trial started with a 1-second pre-fixation period followed by the presentation of 2-second fixation cue at the center of the screen prior to the stimuli onset. Participants were instructed to maintain their focus at the centre of the stimuli after one of the three gratings was presented. They were also asked to either hit the left-click with their index finger when their visual discomforts had reached the maximum (typically 2 to 10 seconds) or the right-click with their middle finger if they did not have any forms of visual discomforts after an 8 second counting in their minds. There was an 7-second inter-stimulus interval followed by the participant’s response before the onset of the fixation of the next trial (see Fig. 1 for the trial sequence). Each type of the stimuli was presented for 50 repetitions in pseudo-random order. A total of 150 trials were separated into 10 blocks with breaks in between. To examine the effect of repeated stimulation, trial 1-75 and trial 76 -150 were further coded as a 2-level independent variable: 1^st^ half and 2^nd^ half, and later be compared in our EEG analyses.

**Fig 1.**
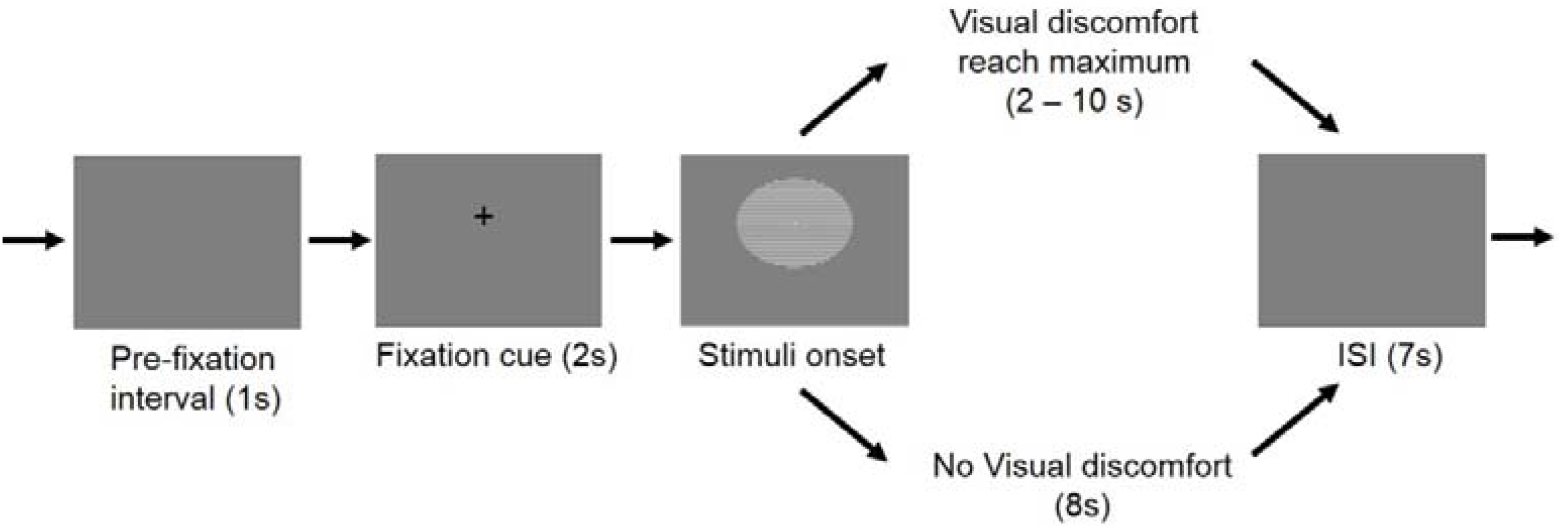
Trial sequence.

### 2.3 EEG recording and preprocessing

The 64-channel EEG signal was recorded at 500 Hz by an EEGO Sports amplifier (ANT Neuro) and Waveguard caps containing Ag/AgCl electrodes in which impedances were kept below 20 kΩ. AFz was used as ground while CPz was used as an on-line reference which was subsequently re-referenced off-line to average reference. Two pairs of bipolar EOG electrodes were used to capture the horizontal (located at the outer canthi of left and right eyes) and vertical (located at the left lid-cheek junction and above left eyebrow) eye movements.

The preprocessing of the data was performed in Matlab using EEGLAB (version 14.1.2b; Delorme and Makeig, 2004). First, the raw data was bandpassed at 0.5 to 40 Hz. The EEG epochs were then locked to the onset (−2 to 3 s) of the visual stimuli. Next, the ocular artefacts (e.g. eye blinks and eye movements) were removed using independent component analysis (ICA). After ICA pruning the data was once again inspected manually and trials with excessive noise rejected. Finally, Trials with responses given in less than 1000 ms were also removed to provide a motor-response free window for post-stimulus analyses.

### 2.4 Oscillatory analysis

The data was then transformed and analysed using the Fieldtrip toolbox (Oostenveld, Fries, Maris & Schoffelen, 2011). The event related activities were first computed by calculating the Time-frequency representations (TFRs) of power for each EEG epoch using sliding Hanning tapers with a 3-cycle time window for each frequency (ΔT = 3/f). The power spectra of the epochs were further divided into the pre-fixation cue interval (−3 to -2 s prior to onset of visual gratings), pre-stimulus (−2 to 0 prior to onset of visual gratings) and post-stimulus (0 to 1 s) intervals. The TFRs of power for the pre-fixation cue and pre-stimulus period was represented in absolute power (μV2) with no baseline being selected while the post-stimulus oscillatory activity was assessed in terms of change in power relative to the mean power in the baseline period -700 to -200 ms before the onset of the visual stimuli (Van Diepen, Cohen, Denys et al., 2015, Mazaheri, Segaert, Olichney et al., 2018).

Non-parametric cluster-based permutation analysis (Maris & Oostenveld, 2007) were conducted on the two intervals separately. In this method, the neighbouring spatiotemporal sample data was clustered if the mean amplitude differences between migraine and control exceeded the threshold at 5% significance level. The electrode-time clusters with a Monte Carlo *p*-value less than 0.025 (two-tailed) was considered as significant (simulated by 5000 iterations), suggesting a between-group statistically difference.

## 3. Results

### Behavioural data

#### Migraine patients exhibited greater discomfort to the visual stimuli

For each participant, the number of trials indicating discomfort (i.e left mouse clicked trials) were divided by the total number of trials, which produced the “fraction of discomforting trials” as the dependent measure for each grating condition. A two-way mixed ANOVA with repeated measure on grating (migraine vs. control x HF vs. MF vs. LF) was conducted. The results revealed there was a significant main effect of migraine, *F* (1, 53) = 11.6, *p* = .001, while the interaction effect was not significant, *F* < 0.2. Due to the unequal group variance and non-sphericity of the data, a non-parametric Friedman’s test was also conducted which gave a consistent result with the above parametric analysis. As post-hoc measures, Welch’s t-tests showed that migraineurs had experienced visual discomfort in more trials compared to control in all three conditions (Fig 2), HF: t = 2.87, *p* = .006, (mean: 82.7% vs. 57.5%); MF: t = 2.95, *p* = .005, (mean: 93.5% vs. 71.0%); LF: t = 2.24, *p* = .029 (mean: 44.5% vs. 23.7%) (all *p*-value remained significant after false-discovery rate correction).

**Fig 2.**
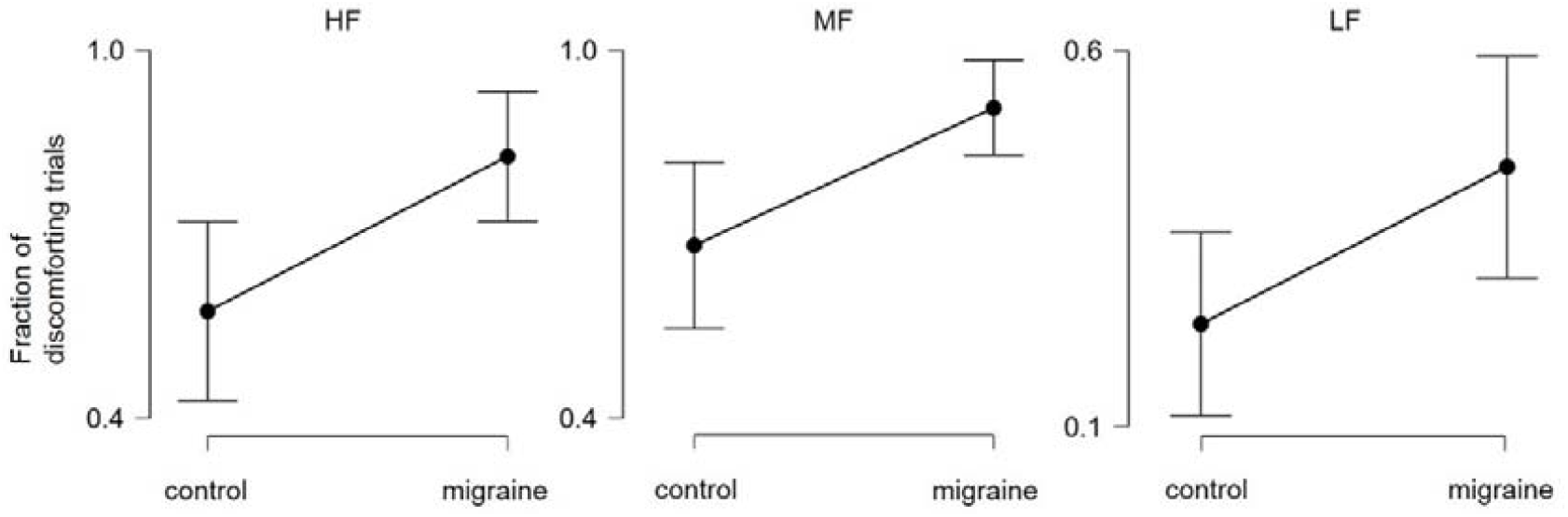
The mean fraction (with 95% CI) of discomforting trials for migraine vs. control across 3 conditions.

To investigate the effect of repeated visual stimulation, the dependent measure “change of fraction of discomforting trials” was calculated by subtracting the “fraction of discomforting trials” of the 1^st^ half trials from the 2^nd^ half trials. Another 2-way mixed ANOVA (migraine vs. control x HF vs. MF vs. LF) was then conducted based on this dependent measure. Despite showing no significant main effect of migraine (*F* < 1), there was a marginally significant interaction effect, *F* (2, 106) = 3.32, *p* = .04. Post-hoc tests revealed that, while not reaching significance, migraineurs did have a trend of experiencing more visual discomfort/distortions in the 2^nd^ half of the trials for the MF condition but not HF and LF, Welch’s t = 2.00, *p* = .051, (Fig.3).

**Fig 3.**
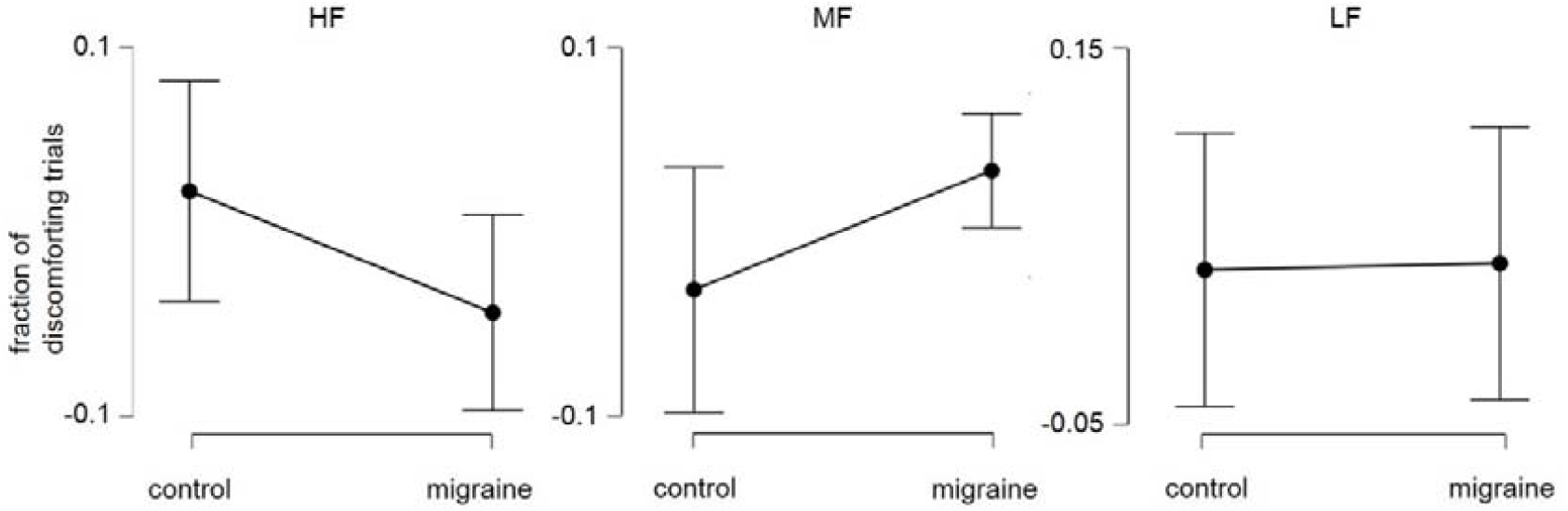
The mean change (2^nd^ half – 1^st^ half) of fraction (with 95% CI) of discomforting trials for migraine vs. control across 3 conditions.

### EEG data

#### Migraine patients had reduced alpha power relative to controls prior to onset of visual grating

Although the main focus of the present study was the induced power changes in the alpha band (8 – 12 Hz), the oscillatory activities in theta (4–7 Hz) and beta (15–20 Hz) band were also examined. The frequency ranges of these bands were chosen and motivated according to prior studies (Klimesch, Doppelmayr, Pachinger et al., 1997; Rommers, Dickson Norton et al., 2017; Sauseng, Klimesch, Freunberger et al., 2006; Mazaheri, Segaert, Olichney et al., 2018; Addante, Watrous, Yonelinas et al., 2011).

We did not observe any significant differences in theta, alpha and beta power between migraineurs and controls in the pre-fixation cue interval. We did however find that post-fixation cue, in the -1.6 to 0.2 s interval relative to the onset of the visual gratings, alpha activity was significantly lower in the migraine patients relative to controls (*p* = .013; Fig. 4). The effect was most pronounced over the occipital-parietal area. While the migraine sufferers and controls did not markedly differ in their baseline level of alpha activity, the significantly reduce alpha activity prior to the onset of the visual grating showed that their visual cortex is in a more excited state prior to the onset of the visual gratings (Fig. 5).

**Fig. 4.**
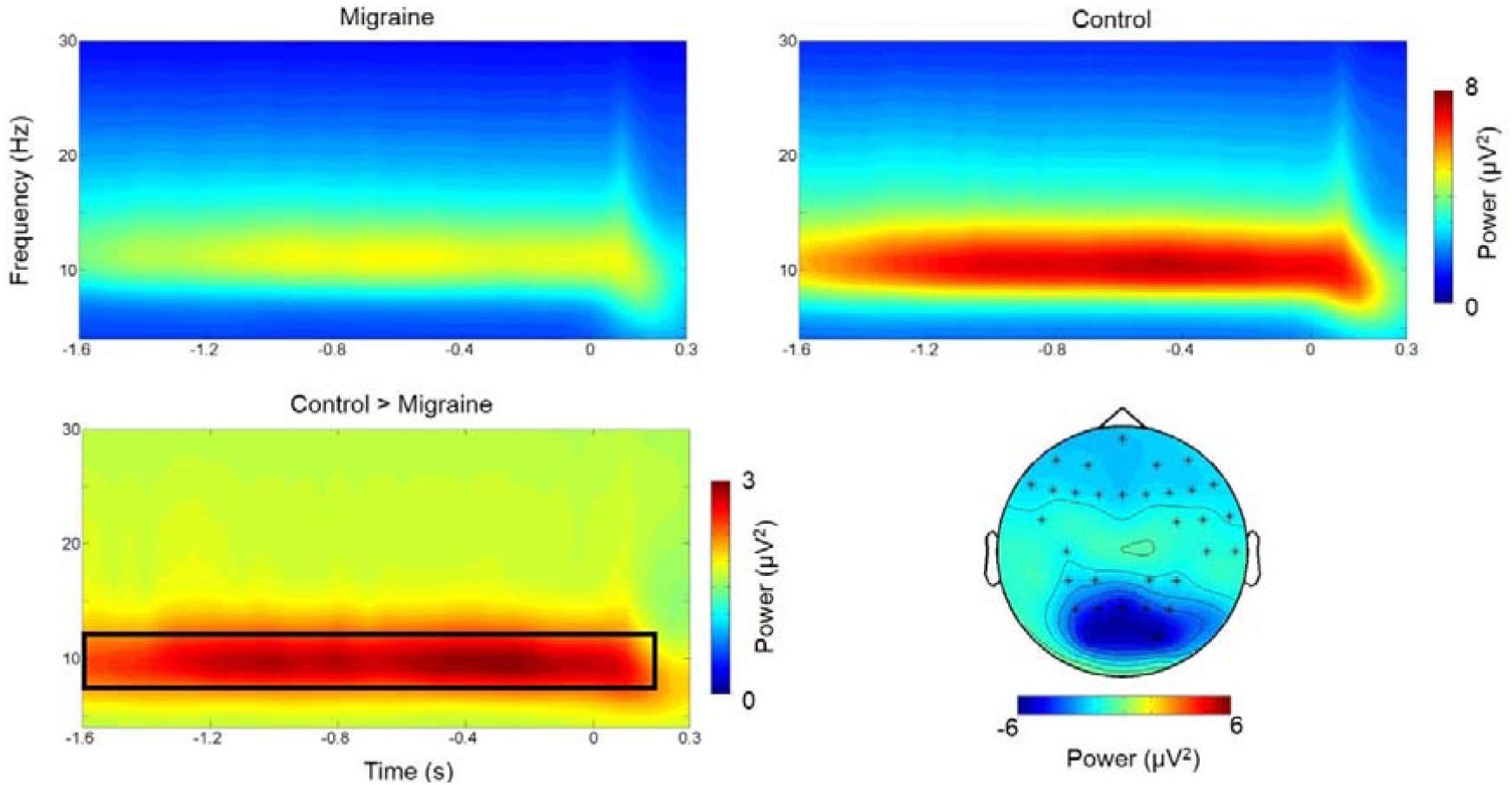
Grand mean (collapsed across all electrodes) time-frequency representation of power and topography of the alpha-band power differences (migraine - control) for the highlighted interval. The electrodes with the maximum effect over the period [-1.6 0.2] were highlighted with *.

**Fig 5.**
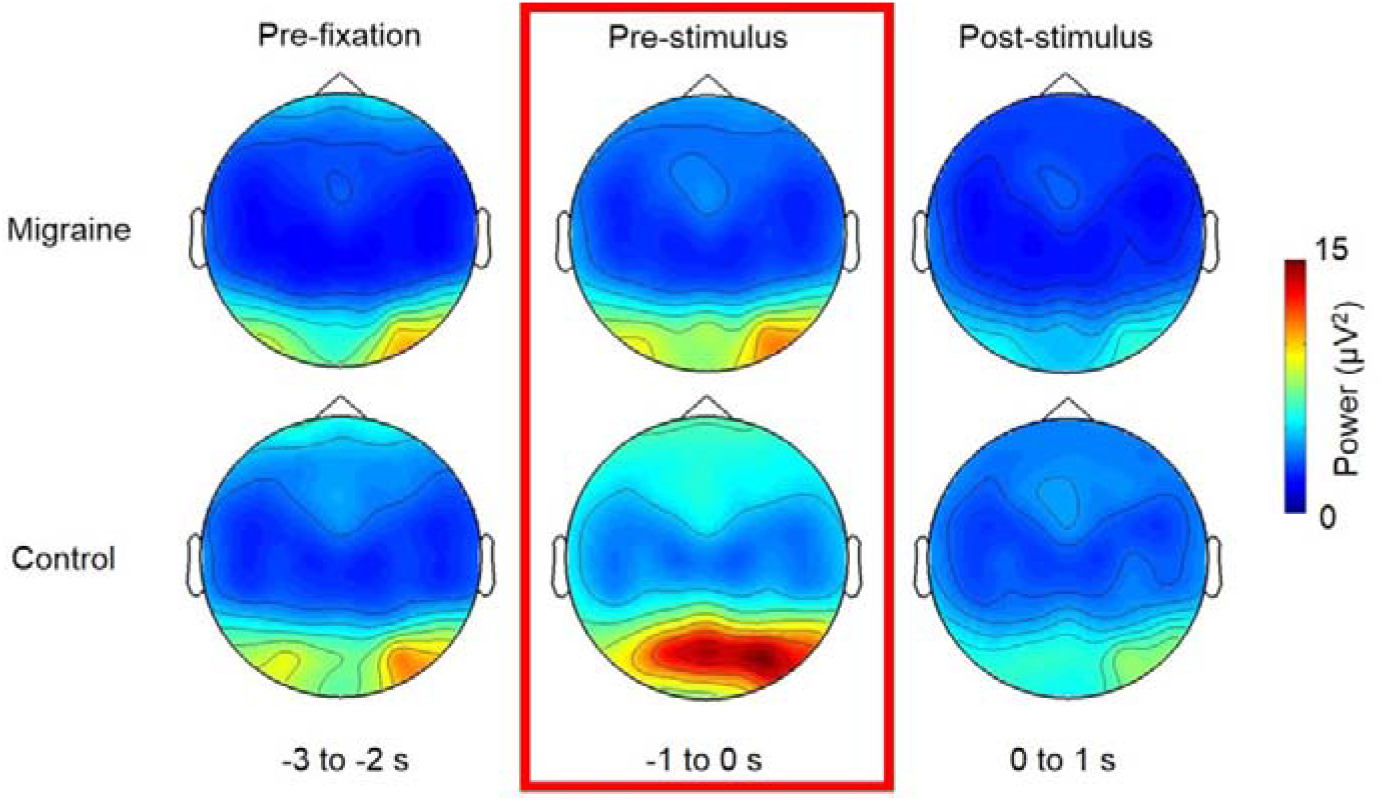
Topographies of the alpha power for pre-fixation, pre-stimulus and post-stimulus period (collapsed 3 grating conditions). The pre-stimulus alpha for migraineurs were significantly lower than controls at the occipital-parietal region, suggesting a more excitable visual-associated cortex.

### For both the migraineurs and controls, pre-stimulus Alpha power was greater in the 2^nd>^ half of the experiment

Next, in order to examine the effect of prolonged visual stimulation, we compared the pre-fixation cue alpha power and pre-stimulus alpha power in the first 75 trials of the experiment relative to trials 76 to 150.

We found that both of the migraine group and control group had significant increase of pre-fixation (migraine: *p* < .001; control: *p* = 004; Fig 6) and pre-stimulus alpha-band power in the 2^nd^ half of the experiment (migraine: *p* < .001; control: *p* < 001; Fig 7). However, the magnitude of increase was not significantly different between migraine and control groups for both pre-fixation and pre-stimulus interval (Fig 6D & 7D). We observed that the pre-stimulus alpha power was consistently lower in the migraineurs (Fig 7C) for both 1^st^ and 2^nd^ half of the experiment (1^st^ half, *p* = .013, 2^nd^ half *p* = .019).

**Fig 6.**
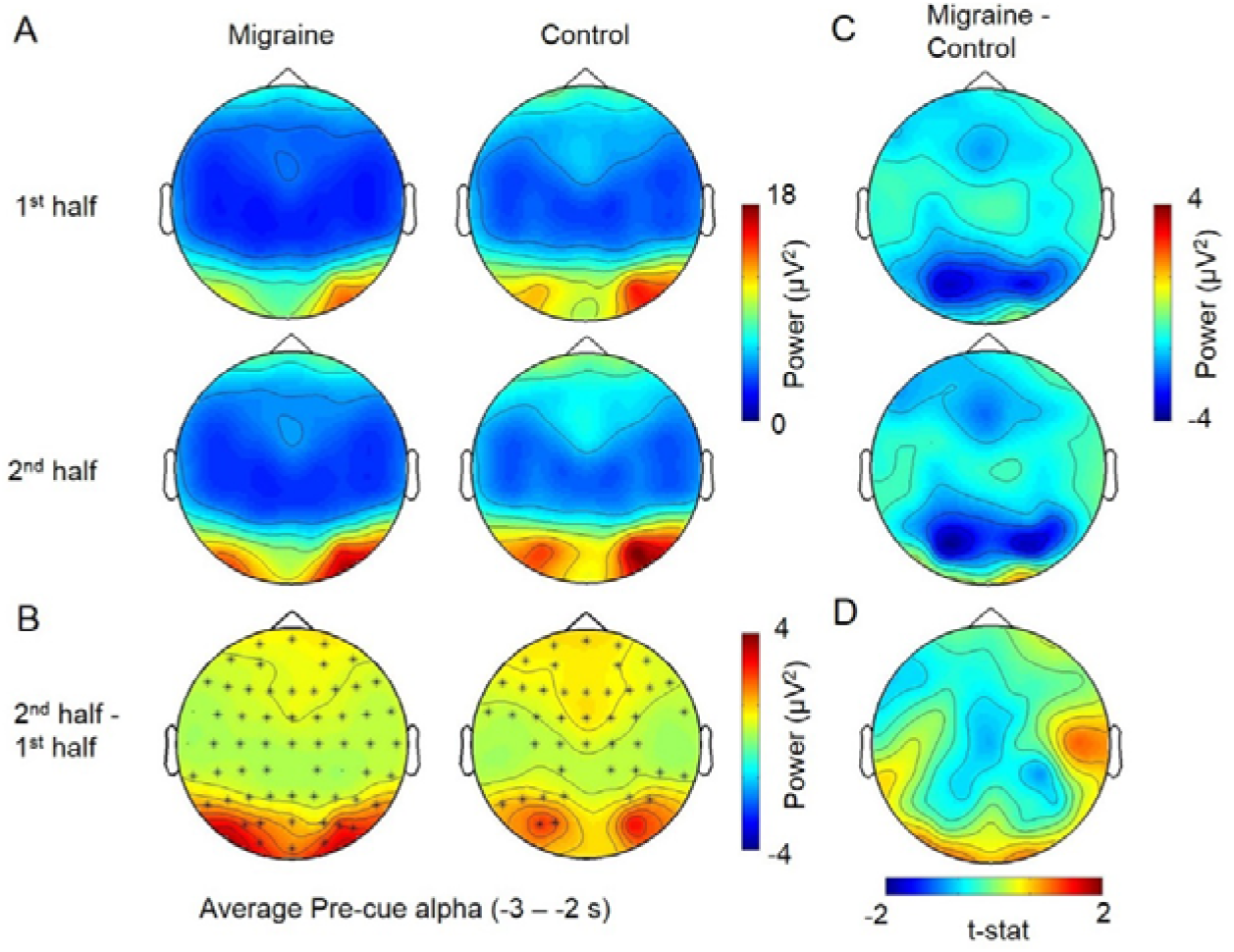
The average pre-fixation alpha power change between 1^st^ half and 2^nd^ half of the experiment. A. The voltage map showed that the pre-cue alpha was the strongest at the occipital area. B. Cluster-based permutation analysis on the pre-fixation interval (−3 to -2s relative to the onset of stimuli) revealed an enhanced alpha power in 2^nd^ half of the experiment for both migraine and controls. The power differences were maximum at the occipital regions (significant channels are highlighted with an asterisk (*) (migraine: *p* < .001; control: *p* = 004). C. There was no significant difference in pre-fixation alpha between migraine and control for both 1^st^ half and 2^nd^ half of the experiment. D. The alpha power increase in the 2^nd^ half were also not significantly different between migraine and control.

**Fig 7.**
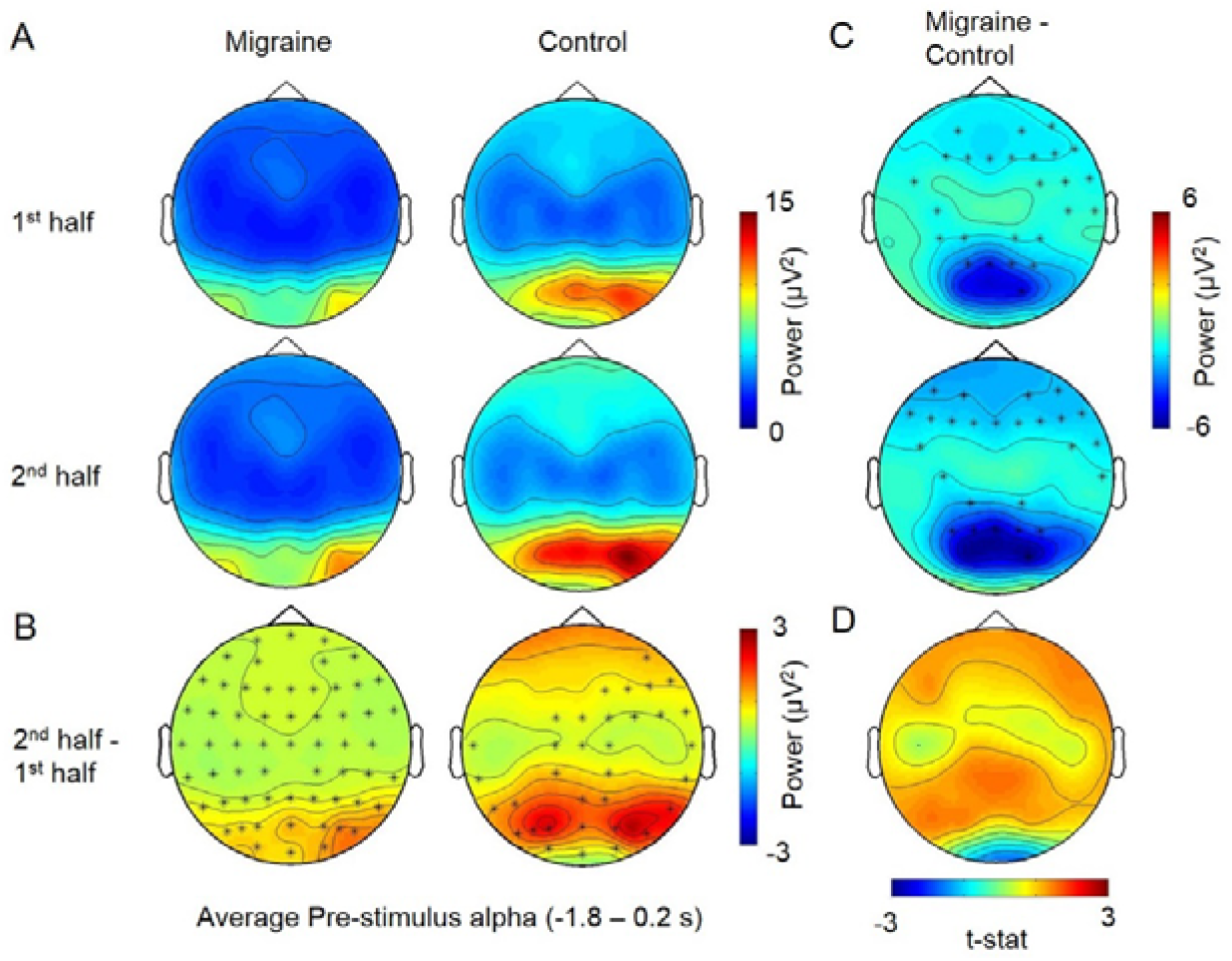
The average pre-stimulus alpha power change between 1^st^ half and 2^nd^ half of the experiment. A. The voltage map showed that the pre-fixation alpha was the strongest at the occipital area. B. Cluster-based permutation analyses displayed one significant cluster for migraine (*p* = 0.0002, t = -1.8 to 0.25) and two for control (*p* = 0.0008, t = -1.8 to -0.75; *p* = 0.002, t = -0.7 to 0.2). The significant channels are highlighted by *. C. For the between group differences, there were one significant cluster for 1^st^ half (*p* = .013) and one for 2^nd^ half (*p* = .019) at the [-1.8 to 0.2] interval, with the alpha power differences mainly distributed over the parietal-occipital region. D. The pre-stimulus alpha power increase in the 2^nd^ half were also not significantly different between migraine and control groups.

### No difference in post-stimulus alpha suppression between migraineurs and match controls

The visual stimuli induced a theta power (4-7 Hz) increase peaking at around 200 ms after the stimulus onset in both migraine and control group across all three experimental conditions (HF, MF and LF; Fig 8). There were also alpha and beta power decreases starting at 300 ms after the grating onset in all conditions. The cluster-based permutation analyses at the post-stimulus interval (0 – 1 s) did not find any significant differences between migraine and control in terms of the magnitude of alpha and beta power decrease and theta power increase across all conditions, all monte-carlo *p* > 0.05.

**Fig 8.**
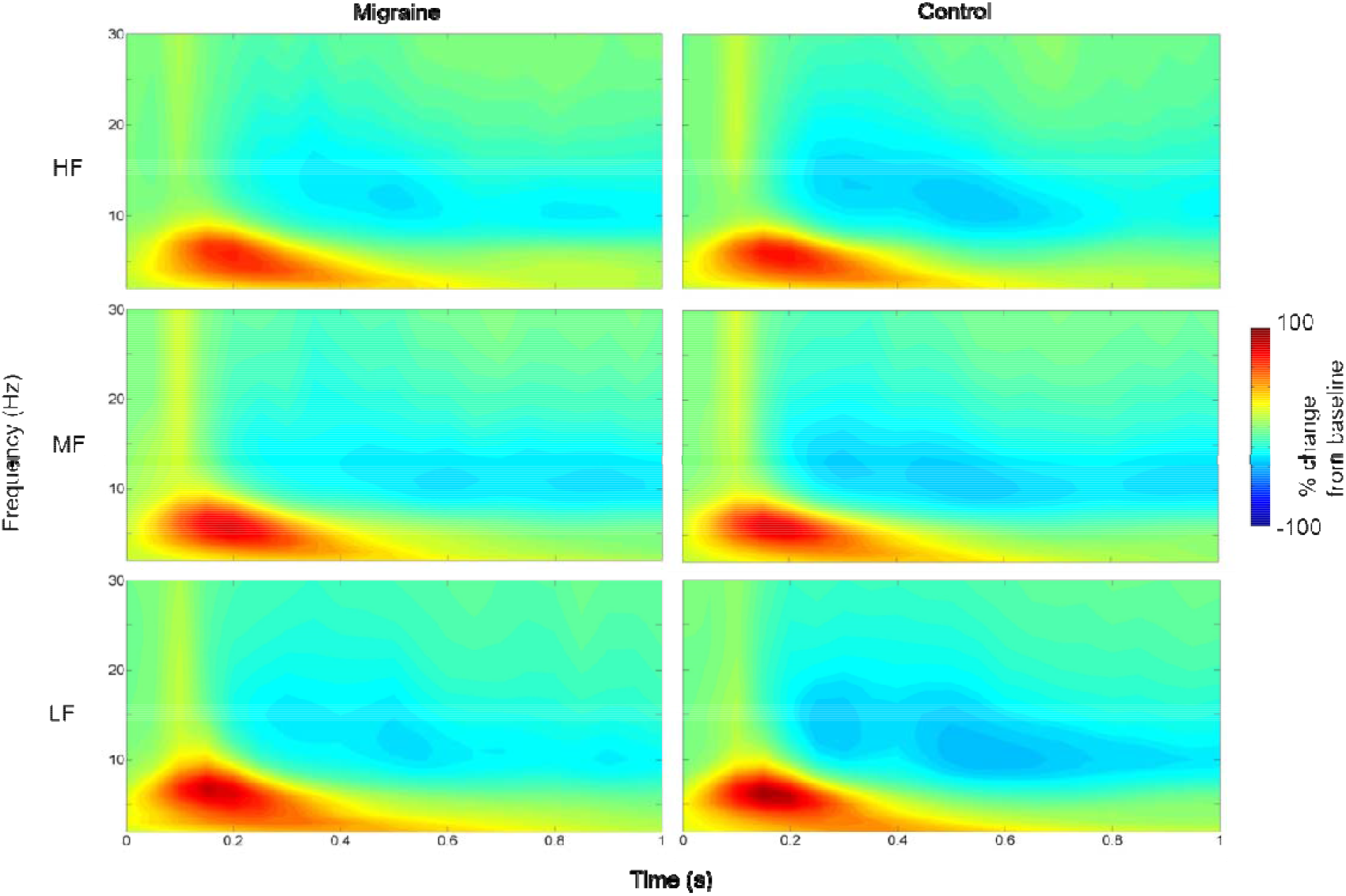
The percentage change of power (migraine vs. control) across the three experiment conditions (HF, MF and LF). The spectrogram indicated the percentage change of power using the pre-stimulus interval -700 to -200 ms before the stimulus onset as the baseline.

### Prolonged visual stimulation enhanced post-stimulus alpha suppression to MF gratings in Migraineurs but not controls

Finally, we examined the effect of prolonged visual stimulation on post-stimulus alpha modulation. We first focused our analysis on the alpha suppression to the MF grating given that it was the grating reported to be causing the most visual discomfort.

We found that for the migraine patients, alpha suppression to the MF grating was significantly greater in 2^nd^ half of the experiment 350 ms to 700 ms (*p* = 0.024) after the MF grating onset (Fig 9A & 9B.). This enhanced suppression was maximal over the central-parietal electrodes. On the other hand, the post-stimulus alpha suppression to MF grating for controls was not significantly different between the 1^st^ half and 2^nd^ half of the experiment.

**Fig 9.**
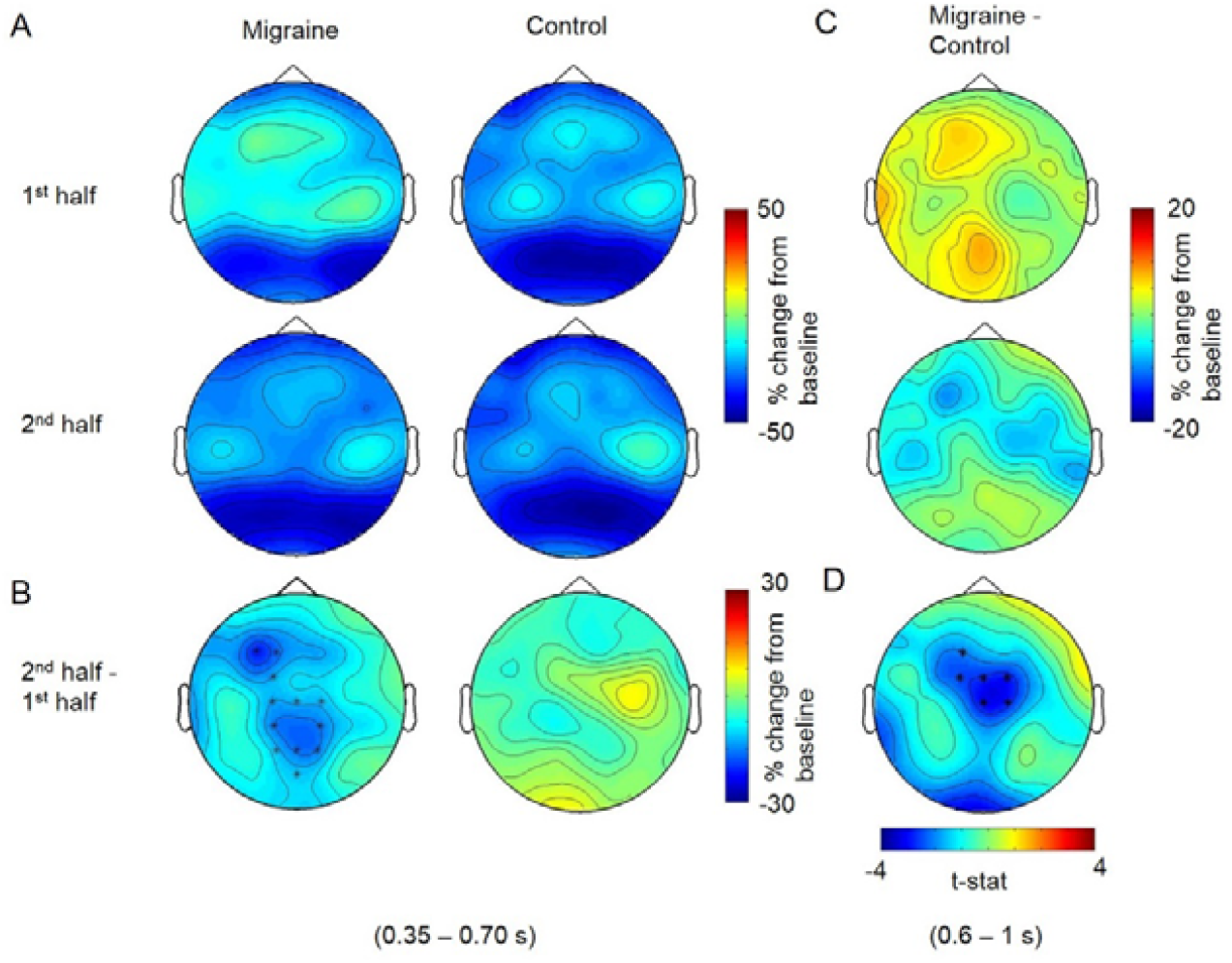
The average post-stimulus alpha power change between 1^st^ half and 2^nd^ half of the experiment for MF condition. A. Cluster-based permutation analysis on the post-stimulus interval (0 to 1s after the MF grating onset) revealed an enhanced alpha suppression in 2^nd^ half of the experiment for migraine between 350 – 700 ms after the stimulus onset. B. The significant channels (highlighted with *) were distributed around central parietal regions. C. There was no significant difference in alpha suppression between migraine and control for both 1^st^ half and 2^nd^ half of the experiment. D. The average alpha suppression (600 – 1000 ms after the stimulus onset) for migraine was stronger in the 2^nd^ half around the central-frontal region. The significant cluster (highlighted with ^*^) indicated the maximum differences in alpha suppression between migraine and control.

To further evaluate the interaction effect of migraine and repeated stimulation on alpha suppression, the alpha power change of 2^nd^ half was subtracted from the 1^st^ half separately for migraine and the control. The resultant data was then subjected to another cluster-based permutation analysis to obtain the between-group effect (migraine vs. control). The result revealed a marginally significant cluster with the effect maximally distributed at the central areas, *p* = .036 around 0.6 to 1 s after the MF grating onset, indicating a stronger alpha suppression in the 2^nd^ half of the study for migraine.

We repeated the above analyses for the HF and LF gratings, however we did not observe a significant difference in alpha suppression between 2^nd^ and 1^st^ half of the experiment neither in the migraineurs nor controls. We also conducted the same analyses for theta and beta activity, which also did not yield any significant differences between migraineurs and controls.

## 4. Discussion

In the present study, we used modulation of the ongoing alpha activity induced by the onset of visual stimuli to assess the excitability of the visual cortex of migraine patients in anticipation of a visual grating, as well as during its processing. We also examined how the alpha modulation during these periods changed from the 1^st^ half of the experiment, to 2^nd^ half, allowing us to assess the impact of prolonged visual stimulation. We focused our investigation on modulations of oscillatory activity in the alpha range given that previous work has found support of this rhythm to be involved in gating of visual input (Jensen & Mazaheri, 2010). There were no significant differences in baseline alpha power between the migraineurs and non-migraine controls. In contrast, alpha power was reliably reduced for migraineurs in the pre-stimulus period prior to the expected onset of the visual gratings.

We did not observe an overall difference in the post-stimulus suppression of alpha activity between the migraineurs and non-migraineur. However, we did observe that migraineurs had significantly more alpha suppression to the visual grating associated with the greatest visual discomfort (MF grating), in the 2^nd^ half of the experiment. We interpret the lower pre-stimulus alpha power seen in migraineurs to reflect that their visual cortex is in a more excitable state in anticipation of the arrival of the visual stimuli via a perceptual learning mechanism. Moreover, the increased alpha suppression to the MF observed in this group suggests that their visual cortex is becoming hyper-sensitised to repeated stimulation rather than desensitised. We will now discuss these findings in greater detail.

### 4.1. Alpha power in pre-stimulus period

Migraineurs persistently showed a pre-stimulus alpha power deficit maximally covering the occipital regions of the brain, During the pre-stimulus period, participants were required to maintain their vision on a steady fixation point, which functioned as a visual cue to hint the onset of the visual stimuli. With the current setup, the visual target would always appear at the same temporal and spatial position which made the stimulus onset being completely predictable. Such a dominance of alpha-band oscillations in the pre-stimulus interval was expected and also in-line with previous literature in which alpha rhythm were found to predict visual detection (Busch, Dubois, & VanRullen, 2009), discrimination (van Dijk, Schoffelen, Oostenveld et al., 2008), awareness (Mathewson, Gratton, Fabiani et al., 2009) and the induction of phosphenes (Dugue, Marque, & VanRullen, 2011). In these visual experiments, researchers found that the phase angle and power of alpha oscillations preceding the missed or detected visual targets were significantly different (Samaha, Bauer, Cimaroli et al., 2015; Busch, Dubois, & VanRullen, 2009). As a result, some researchers proposed that the sensory system was modulated to an ideal “excitability state” by top-down temporal prediction and therefore, alpha-band oscillations might indicate the “excitability state” of the sensory system. Functionally speaking, alpha oscillations might activate the local inhibitory neurons at the visual cortex in order to suppress/filter excessive visual input (van Kerkoerle, Self, Dagnino et al., 2014; Olsen, Bortone, Adesnik et al., 2012; Clayton, Yeung, & Kadosh, 2018). Therefore, with diminished alpha-band activities, more sensory neurons might be activated. Additionally, pre-stimulus occipital alpha was found to indicate the enhanced excitability of the visual cortex (Lange, Oostenveld, & Fries, 2013).

Interestingly, in a more demanding visual detection task (e.g. Busch, Dubois, & VanRullen, 2009; Ergenoglu, Demiralp, Bayraktaroglu et al., 2004; Bauer, Stenner, Friston et al., 2014), the detection of a target was associated with a decrease in pre-stimulus alpha-band power. In our study, we used a non-cognitive demanding task together with aversive stimuli, where pre-stimulus alpha-power was instead tuned to a higher level. We speculated that such an increment of alpha power could re-adjust the sensory cortex into a suitable excitability state after the higher cortical area eventually learnt that the stimuli were irritating. We suggested that such a top-down guided perceptual learning process (Ahissar & Hochstein, 2004) was beneficial to the participants since inactivating the sensory system might relieve the discomforting sensation brought by the visual stimulation from the gratings.

Based on the behavioural data, migraineurs manifested more visual discomfort in response to all types of gratings. An intact alpha modulated neuronal circuit should exhibit an inhibited excitability state. Therefore, it was a clear evidence showing that migraineurs had an impairment with a lower ceiling of alpha-band power (i.e. more hyperexcitable) despite knowing that the upcoming visual stimulation would be aversive. With alpha power known to be associated with the peak amplitude of event-related potential components, for example, P1 (Fellinger, Klimesch, Gruber et al., 2011), N1-P2 (Brandt & Jansen. 1991) and P3 (Ergenoglu, Demiralp, Bayraktaroglu et al., 2004), the reduced pre-stimulus alpha on migraineurs in this experiment could explain their abnormal increased of VEP components found in the early and recent literatures (Diener, Ndosi, Koletzki et al., 1985; Sand, Zhitniy, White et al., 2008; Fong et al., 2020; Oelkers, Grosser, Lang et al., 1999). Additionally, as we suggested, a lower peak alpha might influence the visual perceptual learning process, which could be associated with the poorer performance of migraine patients in certain visual tasks where they must learn to suppress the visual noise in order to perform (Wagner, Manahilov, Loffler et al., 2010; Tibber, Kelly, Jansari et al., 2014). Another symptom of migraine -photosensitivity was also linked to decreased posterior alpha activities (Vaudano, Ruggieri, Avanzini et al., 2017). Collectively, our studies together with the findings in previous literatures all supported the notion of cortical hyperexcitability of migraineurs, which could be driven by the dysfunction of alpha-band activity regulation.

### 4.2. Sensitisation specifically to grating in medium frequency

In the current experiment, participants had to make a behavioural response in every trial in respect of their visual experiences. Though any main effect of migraine on the extent of alpha suppression was lacking, both groups demonstrated stronger cortical activations probably due to increase in visual gain or spatial attention starting from 400 ms after the stimulus onset (Rihs, Michel & Thut, 2007; Peterson & Voytek, 2017). Such reduction of alpha might also be associated with a local change of cerebral metabolic rate (Cook, O’Hara, Uijtdehaage et al., 1998). As the experiment progressed, there was a general increase of baseline alpha power for both migraine and controls. Without any significant group differences, such effect might not be associated with migraine pathology, therefore, it was not the main focus of the present study (perhaps due to fatigue; see Boksem, Meijman, & Lorist, 2005).

Apart from this, we observed that the alpha suppression/sensitisation was maximally localised at the occipital region (see Fig 9A). More interestingly, migraine patients displayed a strengthened alpha suppression specifically to grating in medium frequency (3 cpd) by repeated stimulations. These findings are new and have not been reported in the literature previously. Here we make a few tentative suggestions. First, it could be indirectly caused by cognitive fatigue rather than sensitisation. Mathematically speaking, when there was an increase in baseline and pre-stimulus alpha, a similar/unchanged level of post-stimulus alpha and cortical activations at the 2^nd^ half of the experiment would appear as a stronger alpha suppression relatively. However, we did not observe the same effect over the control group and the other experimental conditions (HF & LF), thus, it is not likely that the present finding was mainly driven by a kind of knock-on effect. Another possibility was that the “stronger” alpha suppression of 2^nd^ half trials was indeed a disguise of the “weaker” alpha suppression in the 1^st^ half. In other words, such phenomenon indicated a recovery of alpha suppression of the migraine sufferers. However, if the alpha suppression in the 2^nd^ half represented a “back to normal” excitability state for migraineurs, we would expect to see a decrease in visual discomfort rather than increase. Therefore, we believed that the diminished alpha suppression in the 1^st^ half was the “better” state for migraineurs in which the excitability was selectively suppressed to reduce the aversive effect of the MF grating. Such sensory process could be a similar perceptual learning mechanism we discussed in 4.1., which was disrupted in the 2^nd^ half of the experiment due to repeated visual stimulations leading to the elevation of alpha suppression. Since migraineurs also reported to experience visual discomfort in more trials in 2^nd^ half, this hypothesis manifested a better coherence to both the behavioural and electrophysiological data.

Additionally, it is consistent with the long-term “potentiation” phenomenon found in migraine. Potentiation highlighted the improper perceptual learning process accompanied by neuroplasticity, where repeated visual stimulations resulted in enhancement of sensory responses (Sale, Pasquale, Bonaccorsi et al., 2011; Dilekoz, Houben, Eikermann-Haerter et al., 2015). This characteristic, which is in contrary to the habituation effect found in the healthy population, can be commonly seen in migraineurs and often reflected by an enhanced rather than a reduced VEP (Afra, Cecchini, Pasqua et al., 1998; Schoenen, Wang, Albert et al., 1995; Ambrosini, Rossi, Pasqua et al., 2003; Kalita, Bhoi, & Misra, 2014). Habituation, which was widely accepted as an adaptive cortical mechanism mediated by GABAergic inhibitory interneurons (Ramaswami, 2014; Giovannini, Rakovska, benton et al., 2001), was proposed to prevent the sensory cortex from overstimulation (Thompson, Berry, Rinaldi et al., 1979) and lactate accumulation (Sappey-Marinier, Calabrese, Fein et al., 1992). Grating (especially in medium frequency) might stimulate a relatively localised nerve network of the primary visual cortex (see a review of pattern glare: Evans & Stevenson, 2008), thus, the long-term exposure to the MF grating might overload the synthesis or reuptake of the inhibitory neurotransmitter of the impaired GABAergic system of migraineurs.

### 4.3. Limitation and Future direction

The impairment of GABAergic mediated inhibitory network on migraine has been widely discussed in previous literatures (Aurora, Al-Sayeed, & Welch, 1999; Brighina, Palermo, & Fierro, 2009). Apart from visual disturbances, a dysfunctional GABAergic system, is more susceptible to enhanced synaptic transmission, spreading depression (Dilekoz, Houben, Eikermann-Haerter et al., 2015) and the activations of trigeminal nociceptive neurons are all possible cause of the infamous head pain of migraine attack (Cornelison, Woodman, & Durham, 2020; Knight, Bartsch, Kaube et al., 2002). Although migraine research in GABAergic pathway facilitates prophylactic medication development targeting GABA receptors (Sprenger, Viana, & Tassorelli, 2018; Cutrer & Moskowitz, 1996), some research showed that the concentration of GABA between migraineurs and healthy controls were not fundamentally different (Chan, Pitchaimuthu, Wu et al., 2019). In addition, serotonin appeared to be associated with the above network since the treatment of anti-reuptake agent of serotonin was able to restore the function of habituation (Ozkul & Bozlar, 2002). Being the most abundant interneurons of the cerebral cortex, GABAergic interneuron was also known to associate with most cognitive function but not uniquely to the pathology of migraine. Therefore, a deeper investigation at functional, anatomical and even cellular level of GABAergic interneuron might be critical to understand the cause of migraine in the future.

It should be noted that visual disturbances, elevated cortical hyperexcitability and alpha-power deficit by themselves likely cannot provide a full picture of migraine pathophysiology. Moreover, the origin of the alpha-power deficit and how it is associated with the neuropathology of migraine is unknown. Nonetheless, the maximum effect of alpha power differences we found was around the parietal areas rather than localising at the occipital regions which directly received the sensory input from the early visual pathway. It is rather not surprising since recent studies had already shown that the occipital alpha and the excitability of the visual cortex could be modulated by both cortico-cortical (e.g. prefrontal, parietal) and thalamocortical interactions (Helfrich, Huang, Wilson et al., 2017; Lorincz, Kekesi, Juhasz et al., 2009). Recent development on predictive coding and perceptual learning also challenged the idea of perception being a bottom-up process, but a bi-directional and hierarchical integration of information from both the higher-order cortical area and lower-order subcortical area (see Fong, Law, Uka et al., 2020; Rao & Ballard, 1999). In this sense, an abnormality of visual sensations such as visual disturbances do not necessarily equal to a damage at the visual cortex, any inter-connected network could all contribute to such sensory impairment.

In conclusion, our study revealed that migraine patients had pre-stimulus alpha deficit during the anticipation of the visual stimulation. They also manifested sensitisation (increased alpha suppression) selectively to the grating with spatial frequency in 3 cpd by repeated stimulation. With alpha-band oscillation known to be an indicator and mediator of the excitability state, the present study demonstrated the elevated cortical hyperexcitability of migraine. We speculated that it could be the consequence of improper perceptual learning process driven by the dysfunction of GABAergic inhibitory mechanism. Taken together, our study showed converging behavioural and electrophysiological evidence for the cortical hyperexcitability of migraine sufferers which facilitated their experience of visual disturbances.

## Data Availability

The data will be made available when it is accepted.

## Acknowledgements

We thank all the participants for taking part in this study. This research did not receive any specific grant from funding agencies in the public, commercial, or not-for-profit sectors.

